# From the COVID-19 Pandemic to the Mental Health of the Philippines: Modelling the Cascade of Disasters using an Influence Diagram

**DOI:** 10.1101/2025.05.14.25327598

**Authors:** Gabriel Lorenzo Santos, Paul James Montecillo, Jesus Emmanuel Sevilleja, Will Jose, Janinalaine Platero, M. Teresa Tuason, C. Dominik Güss, Vena Pearl A. Bongolan

## Abstract

The COVID-19 pandemic had not only physical, but also mental effects due to the anxiety of being infected, necessity of community quarantine, and the big shift in lifestyle over the course of the pandemic. In this study, a Bayesian network was constructed and informed using the responses of 1,605 Filipinos to a survey conducted online during August - September 2021. The main objective of this study is to identify the critical factors that caused the cascade of disasters from the COVID-19 pandemic to a lower state of well-being for the community, by observing how it affected the mental health of the Philippine citizens. To achieve this goal, a Bayesian network was used to assess the state of the community, which was then extended into an influence diagram. Along with expert opinion on possible interventions (decision nodes), we can obtain an optimal set of interventions/decision nodes in order to lift the community’s state of mental well-being. The study aims to identify the proper decisions to make if another pandemic were to occur as well as become a framework for future applications of the Bayesian network into practical fields.

## 1 Introduction

The COVID-19 pandemic resulted in a long standing lockdown, one of the many affected being the Philippines, which, sadly, had one of the longest if not the longest and most strict quarantines in the world. The pandemic caused a lot of mental stress due to the interruption of day-to-day activities, with uncertainty of the future due to financial troubles and fears over the health of family and friends. Tuason et al. (2023) studied the benefits and costs that COVID-19 brought to the Philippines, looking for their correlation to an individual’s mental health, identified the predictors of the psychological well-being (PWB) of Filipinos during COVID-19 using a survey with 40 questions, obtaining data from variables such as psychological well-being, biological variables (gender, age, and physical health), psychological variables (spirituality, emotional loneliness, social loneliness, and sense of agency), and socio-economic variables (hours communicating on social media, and neighborhood safety, employment security, and income). Although Tuason et al., (2023) brought to light how COVID-19 can affect mental health, the study is limited to looking into the correlation between isolated factors and mental health. In reality, a change in one factor can influence another, leading to conditional dependence.^1^

Bayesian networks are used in working with conditional dependence and unexpected relationships. Barros, et al. (2020) used Bayesian networks in their study to apply conditional dependencies between different states that patients could experience and how these could result in suicidal behavior. The study shows how Bayesian networks can be used to identify key factors that affect a target result but no optimizations were made; no attempt was made to obtain the best-case scenario for interventions.^2^ A solution would be to extend the Bayesian network to use influence diagrams, testing various use-cases to obtain the optimal set of interventions to apply.

This study aims to identify the critical factors that form the cascade of disasters from the COVID-19 pandemic and how it affects mental health using a Bayesian network. It also aims to extend the Bayesian network into an influence diagram in order to identify which interventions are optimal in order to improve mental health. The study uses data from the Philippines and aims to serve as a framework for future mental health studies that require the inclusion of conditional dependence between various factors. We end with a list of recommendations to improve mental health in such a critical period.

## 2 Literature

### 2.1 COVID-19 Pandemic and Mental Health

According to the World Health Organization (2022), the prevalence of anxiety and depression worldwide alarmingly increased by 25%. It was also cited that “one major explanation for the increase is the unprecedented stress caused by the social isolation resulting from the pandemic. Linked to this were constraints on people’s ability to work, seek support from loved ones and engage in their communities. Loneliness, fear of infection, suffering and death for oneself and for loved ones, grief after bereavement and financial worries have also all been cited as stressors leading to anxiety and depression. Among health workers, exhaustion has been a major trigger for suicidal thinking.”^3^ This shows how the root cause of mental health issues during a pandemic is multifaceted. Multiple events that are a result of the pandemic can also cause stress, anxiety, and depression.

Being infected by the virus has a direct effect on people’s mental health. According to Bo et al., (2020 as cited by Vindegaard & Benros, 2020) 96·2% of 714 patients that were hospitalized but were stable experienced post-traumatic stress symptoms.^4^ In another study by Zhang et al. (2020 as cited by Vindegaard & Benros, 2020), there was an increase in the prevalence of depression among the infected patients which is in accordance with the report of WHO (2022).^3,4^

In addition, efforts to contain the virus caused social isolation and loneliness due to limiting access to social relationships (Gonçalves et al., 2020; Hoffart et al., 2020).^5,6^ Hoffart et al. (2020) also suggested that loneliness during the pandemic was exacerbated by rumination and worry.^6^

Multiple research studies show chronic financial strain correlates with higher levels of depressive symptoms across different populations (Aranda & Lincoln, 2011 as cited by Hassan et al., 2021; Zürcher et al., 2019 as cited by Hassan et al., 2021; Tøge, 2016 as cited by Hassan et al., 2021).^7^

### 2.2 Mitigations

Physical activity has been proven to be beneficial for both physical and mental health. In a study done by Zhang et al. (2020) in China, it was discovered that physical activity during the COVID-19 pandemic lessened negative emotions such as depression, anxiety, and stress.^8^ The study of Cruz et al. (2022) in the Philippines also supports this. It was recommended in the study that people do about 108 minutes of light, 80 minutes of moderate, or 45 minutes of vigorous physical activity every day for it to affect their mental health. It is also important to note that this must be done indoors as going to the gym and other public places for physical activity may be a high risk of contracting COVID. During the community quarantine in the Philippines, the physical activity level of university students was seen to decline.^9^ This decline is concerning, as reduced physical activity can lead to both immediate and long-term negative health outcomes, especially during the pandemic. Implementing policies and initiatives that promote physical activity at home would greatly help mental health.

Social distancing and lockdowns were of great importance during the pandemic. However, this resulted in social isolation and loneliness, which have significant impacts on mental health (Loades et al., 2020; Kim et al., 2021; Hoffart et al., 2020).^10,11,6^ Psychological therapy was the most effective form of intervention for loneliness (Williams, 2021).^12^ This is supported by Hamouche (2020), who enumerated mental health services as one of the possible mitigations for mental health during COVID-19. These services may also include mental health hotlines, online consultations, and online courses.^13^ Another mitigation Hoffart et al. (2020) identified was engaging in new positive activities at home. This may serve as a distraction since according to Hoffart et al. (2020), the cause of loneliness during the pandemic may also be rumination and worry.

Job and financial security are also essential factors to consider for mental health during a pandemic, since these two have a significant impact on the mental well-being of people (Hoffart et al., 2020).^6^ Hamouche (2020) enumerated multiple mitigations that could assist employees during the time of pandemic. The importance of teleworking was emphasized as this gives people the opportunity to keep their jobs and maintain financial stability.^13^ The effectiveness of emergency packages in mitigating the financial impact of the pandemic has also been mentioned, but a rather counterintuitive result was noted. The Philippines implemented such measures during the pandemic, such as *ayuda (social amelioration)* packages, and worker protection programs. The government was able to provide financial assistance worth ₱5,000- ₱8,000 and ayuda packages worth ₱1,000-₱4,000 per household (Cho and Johnson, 2022).^14^

Cho and Johnson have specified that due to these emergency packages, families who received the support were less likely to report food insecurity. It was also mentioned in the report that despite these measures, two-thirds of households experienced a decline in per capita income. The poverty rate and hunger rate are still expected to rise significantly due to the consequences of the pandemic, which could also lead to the rise in mental health issues.^14^

## 3 Methodology

This study constructs a Bayesian Network to model and analyze the cascading effects of the COVID-19 pandemic on the mental health of a surveyed population in the Philippines. Furthermore, it extends the network into an Influence Diagram to evaluate the impact of mitigation strategies on the final mental health outcomes.

### 3.1 Bayesian Networks and Influence Diagrams

Bayesian networks (BNs) are probabilistic graphical models that represent the joint probability distribution of random variables and their conditional dependencies using a directed acyclic graph (Sucar, 2015).^15^ The nodes in a BN represent random variables, while directed edges illustrate the probabilistic relationships between these variables. Each node is associated with a conditional probability table that defines the probability of each state of the variable, given its parents in the network (Chen et al., 2024).^16^

Bayesian networks enable inference and prediction under uncertainty because they can model causal relationships between variables (Dong et al., 2019).^17^ This makes them particularly useful in understanding and simulating the cascading and compounding effects between parent-child events (Zhang et al., 2023; Vatteri et al., 2022; Dong et al., 2019).^17–19^ However, constructing the conditional probability tables as parameters to the BN requires large amounts of data, making the development of the BN data-intensive (Dong et al., 2020).^20^ Building the network structure also incorporates expert knowledge, adding subjectivity to the modeling process and requiring the combination of divergent expert judgments (Uusitalo, 2007).^21^

On the other hand, influence diagrams (IDs) extend BNs by incorporating decision and utility nodes, visually representing decision-making under uncertainty (Zhu et al., 2021).^22^ Aside from modeling causal relationships between variables, IDs include the causal effect of the decisions on the variables and calculate its impact to the objective or utilities. The ultimate goal in solving influence diagrams is to determine the optimal strategy (i.e., the set of decisions that maximizes expected utility) and the corresponding optimal utility or value (Shachter & Bhattacharjya, 2011).^23^ This study implements the influence diagram in Figure 1 using the pyAgrum library in Python (Ducamp et al., 2020).^24^

**Figure 1:**
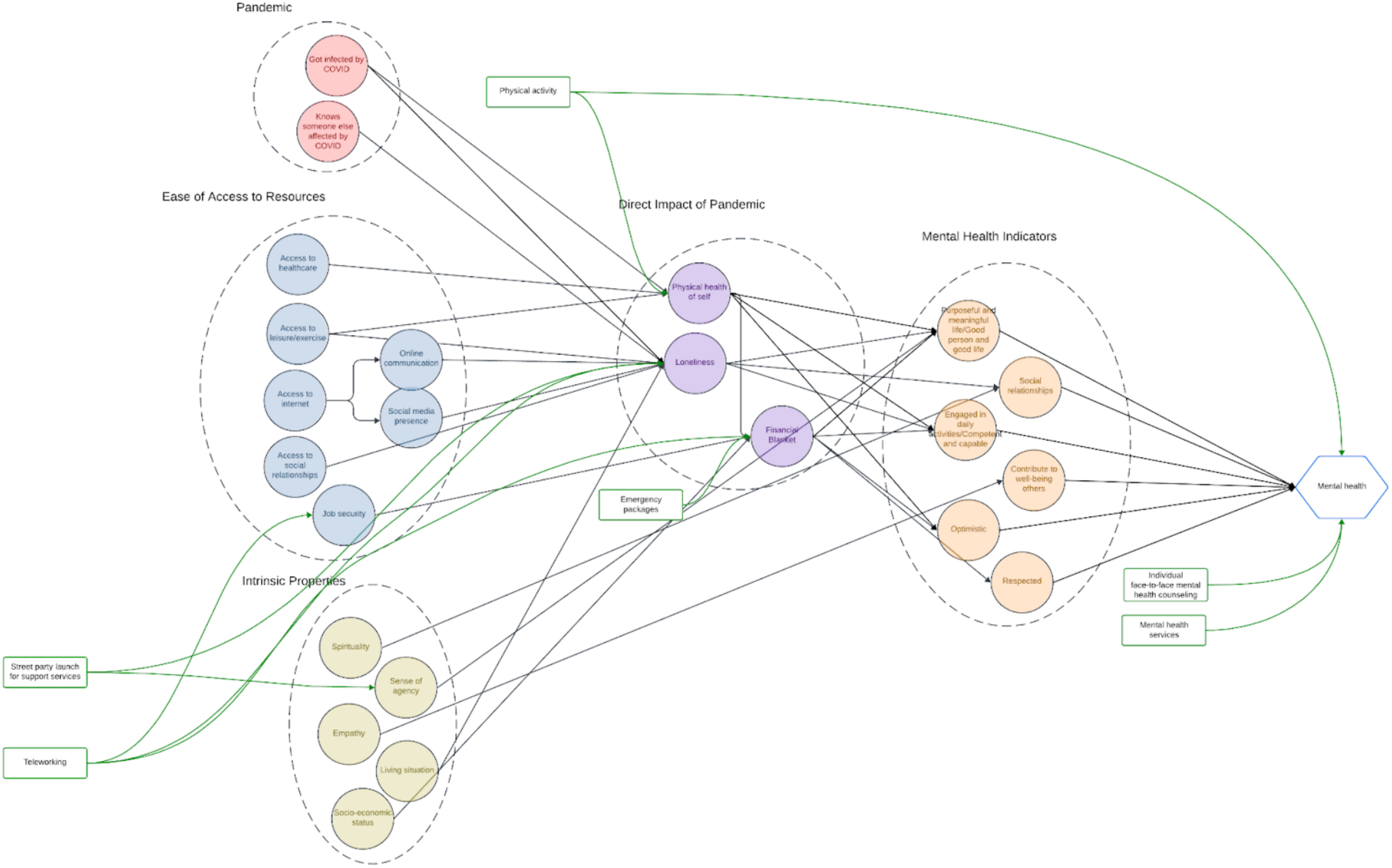
Bayesian influence diagram of the cascade of disasters from COVID-19 to mental health.

### 3.2 Structure of the Influence Diagram

Figure 1 shows the influence diagram used to model and assess the cascading impact of the COVID-19 pandemic on the mental health of Filipinos. The method follows the cascading crisis events Bayesian Network (CCEBN) approach of Qiu et al. (2014), extended with decision and utility nodes to arrive at optimal interventions.^25^ Each event is represented as a single crisis event Bayesian Network, where the event state variables are affected by input chance nodes and decision nodes, representing external environmental variables and control decisions, respectively. Depending on the interaction of the input chance nodes and the decision nodes with the state variables, it results in certain response and loss output variables. Cascade occurs when the prior event response output variables become the input environment variables to the triggered event (Qiu et al., 2014).^25^ Single events are represented by circles with broken outlines in Figure 1.

#### 3.2.1 Chance Nodes

Chance nodes show the probabilities of their state or event. In Figure 1, the variables from the questionnaire of Tuason et al. (2023) are represented by the chance nodes. How each variable influences one another is extracted through expert elicitation with psychiatrists, mental health professionals, and researchers’ practical judgment. The influence of one node on another is represented by arrows in Figure 1.

The flow of events follows a similar structure used by Qiu et al. (2014), arranged in layers, to analyze the components of an event from a systems theory perspective.^25^ The first layer represents the independent crisis or events, in this case, the pandemic itself, an individual’s access to resources, and an individual’s intrinsic properties, which are defined as follows:

- Pandemic — This category defines the pandemic as both a personal and shared experience. The survey questions used to formulate the nodes under this category were whether people got infected by COVID-19 and whether they knew someone else affected by the virus.
- Ease of Access to Resources — The nodes grouped in this category are based on the survey questions measuring how the pandemic affects the accessibility of essential services and resources crucial for the respondents’ well-being and quality of life.
- Intrinsic Properties - The nodes under intrinsic properties were established using the survey questions designed to measure characteristics inherent to individuals, such as the questions for psychological variables.

The second layer represents the direct impact of the pandemic on the individual’s physical and economic capability. This includes physical health of self, loneliness, and financial blanket. Although social and emotional loneliness were separately measured by Tuason et al. (2023), this study combines them since their parent nodes are the same. The financial blanket node was formulated based on how easily households can make ends meet.

The third layer represents an individual’s mental health indicators. The nodes from this group were based on the eight questions in the survey used to assess psychological well-being (Tuason et al., 2023).^1^ Each question forms a node, although some similar questions were merged into a single node for simplicity.

The fourth and final layer represents an individual’s mental health state, modeled in Figure 1 as the utility node being optimized. The basis for the node in this group is the respondents’ overall psychological well-being. This is the final measure to identify the optimal interventions in the overall cascade.

To develop the conditional probability tables for each node, the responses from the survey conducted online by Tuason et al. (2023) with 1,605 Filipinos were used.^1^ Each question is assigned to a chance node. Although all the chance nodes in the influence diagram came from the survey questions, not all survey questions were used to create the network nodes. The responses were standardized for each node and grouped into label states 1 to 3, signifying the degree of response to the questions. For example, responses to the question regarding access to healthcare are grouped into 1 (no access), 2 (limited access), and 3 (full access). Using BNLearner function in pyAgrum (Ducamp et al., 2020), the conditional probabilities are calculated from the final transformed survey data and are stored in each node’s respective conditional probability tables.^24^

#### 3.2.2 Mental Health Treatment and Support as Decision Nodes

Through expert elicitation, mental health experts and psychologists in the research team derived various treatments and support for mental health decline and their efficacies, grounded on research and experience applied in the context of the Filipino community. A review by Tuason and Güss (see Appendix A) indicated mindfulness programs and individual follow-up, door-to-door quick response groups, and healing rituals as effective interventions due to their adherence to the Filipino psychology framework. Hamouche (2020) included mental health services, teleworking, and emergency packages as helpful interventions for the mental health decline of working employees due to the COVID-19 pandemic.^13^ The research team added face-to-face and remote mental health counseling as additional mitigations.

However, a study on general telehealth in the Philippines during the pandemic (Noceda et al., 2023) found both approaches were preferred indecisively.^26^ Individual physical activity was also pushed for its benefits to physical and mental health, as it helped reduce negative emotions during the pandemic (Zhang et al., 2020).^8^ Decision-makers may implement policies and initiatives that promote active lifestyles among the population to alleviate the negative impacts of the pandemic.

Under lockdown or due to preventative measures, teleworking may be enacted to maintain job security and financial stability. Teleworking limited the population’s mobility and thus reduced the chance of COVID-19 infection and allowed people to continue working. In addition, emergency packages that aim to support families financially can address economic stress during the pandemic. These packages can include direct payouts to individuals, loans, and business guarantees, helping people maintain financial security. These measures can contribute to better mental health outcomes by alleviating financial pressures.

Table 1 lists these interventions and their perceived efficacies. These interventions are used as decision nodes in the influence diagram, adding controlling or limiting influence to the crisis or event characteristics. The efficacies, however, are derived from expert opinion, as there can be no accurate way to measure the efficacy of each intervention at this point.

The efficacy values of the different interventions in this study produce a scaling effect on the state probabilities of the chance node they are trying to control. The decision nodes were assigned ‘yes’ and ‘no,’ representing whether the intervention was activated or not, respectively. The efficacy is applied if the decision node is turned ‘yes.’ For example, a particular intervention with a 50% efficacy of improving mental health reduces the likelihood of getting low and medium states by 50%. It thus increases the chances for a high state and, therefore, a better chance of higher mental health states. If the decision node is set to ‘no,’ the original probabilities are retained.

#### 3.2.3 Mental Health State as Utility Node

Given the stressors from the COVID-19 pandemic, its cascading impact, and coping activities, mental health is the primary outcome being observed and optimized in this study, making the mental health state the utility node of the influence diagram. This focus on mental health allows us to measure the extent to which people’s well-being was affected by the pandemic and to identify which preventive activities, or a combination of them, have the most significant impact on the state of the respondents’ mental well-being.

### 3.3 Optimal Decisions

The utility node’s value was determined by the probability of achieving a “high” state given various combinations of events. The optimal choices for decision nodes were based on the maximum expected utility (MEU), as implemented in pyAgrum.

## 4 Results

In Figure 2, the conditional probability tables assigned to each individual node can be seen. These conditional probability tables provide the posterior probability that one belongs to each state at any given node. The conditional probabilities seen in each node are reflective of the distribution of the original data but the probabilities were automatically adjusted based on how much each node factors into the succeeding nodes. This is because the dataset fed into the network serves as the prior probabilities and once the network has learned through all of the provided nodes and states, the probabilities would change depending on the probabilities of both its parents and successors. An example of this change would be how the original dataset contained 6% more with a “Mental Health” of 2 or “average”, but this was distributed into the “low” and “high” states based on the learning patterns of the Bayesian network. This leads to more accurate conditional probability tables and predictors as a result.

**Figure 2:**
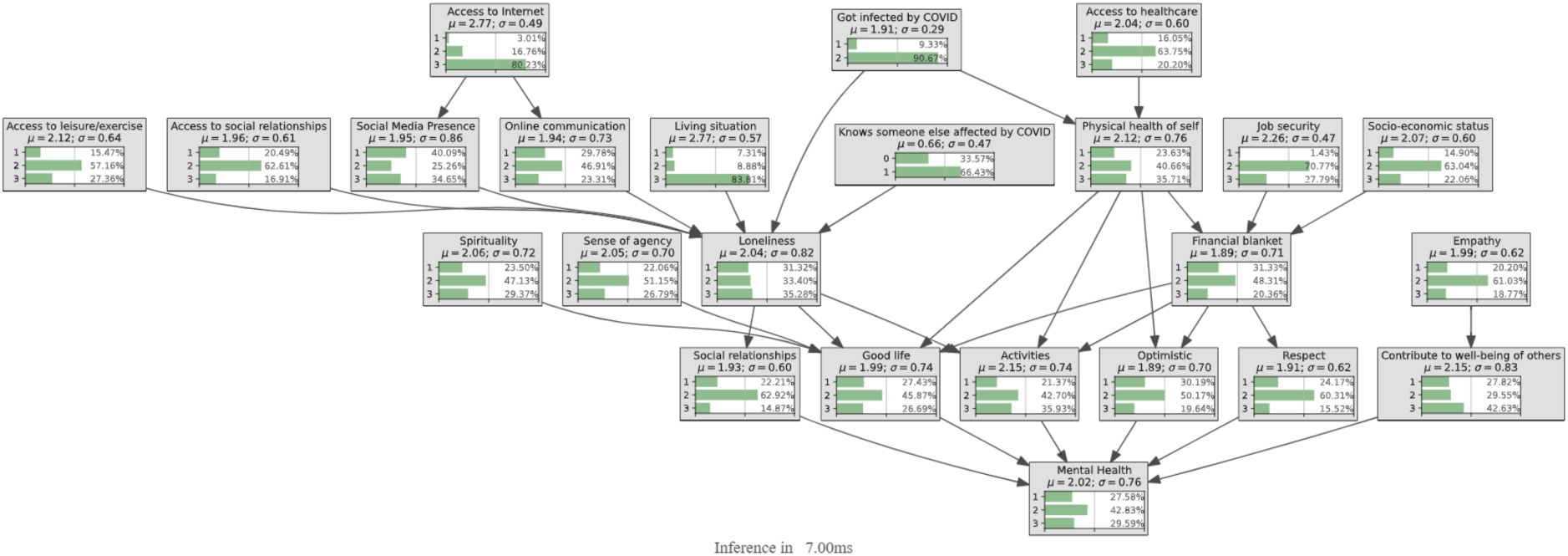
Base conditional probabilities of the Bayesian influence diagram.

Using the Bayesian diagram as a diagnosis tool, one can set the states of the previous nodes and the resulting posterior probability of the “Mental Health” node can change as needed. For example, setting the “Loneliness” of a person to “high”, the change is propagated throughout the network and a new set of probabilities can be obtained. The conditional probability table of “Mental Health” can then be used to see if the person has a “low”, “average”, or “high” mental health state. In looking for optimal interventions, the study uses this concept of manipulating probabilities of the existing Bayesian network as seen in 3.2.

Manipulating the probabilities through the use of interventions, seen in Table 1, shows the results in Table 2 where each scenario is implemented one at a time. If the scenarios were all run at the same time, then it would result in the highest increase in “high” mental health states, but it is assumed that resources are limited and only one intervention can be implemented. As all interventions aim to modify the states to “high”, all of the decisions are seen to have a positive impact since the optimal decision for all states is “yes”. This means that as long as some intervention is made, the overall mental health will increase.

Based on the data presented in Table 2, every decision improves utility. From highest to lowest effect on utility, the nodes are:

- Individual physical activity
- Mindfulness program, individual followup
- Door-to-door quick response group
- Face-to-face counseling
- Remote counseling
- Healing ritual
- Emergency packages
- Teleworking

It can be seen that the order of most effective to least effective interventions is in descending efficacy. This means that the nodes with the highest efficacy, namely Individual physical activity (75% efficacy) and Mindfulness program, individual followup (65% efficacy) are the top two in MEU, while Emergency packages and Teleworking (both 50% efficacy) are the bottom two in MEU. This could be because every intervention directly influences Mental Health, the utility node, which could result in a bias toward efficacy. To test this, we add a weight, *α*, on the efficacy towards the Mental Health node. There are 4 weights of α (0·8, 0·6, 0·4, and 0·2) which will be multiplied only to the efficacy on the utility node.

Based on the data presented in Table 3, the list of interventions from highest to lowest MEU with α = 0·8 is:

- Door-to-door quick response group (from third most effective at α = 1)
- Mindfulness program, individual followup
- Individual physical activity (from most effective at α = 1)
- Face-to-face counseling
- Remote counseling
- Healing ritual
- Emergency packages
- Teleworking

Accordingly, the list of interventions from highest to lowest MEU with α = 0·6 is:

- Door-to-door quick response group
- Mindfulness program, individual followup
- Face-to-face counseling (from fourth most effective at α = 0.8)
- Individual physical activity (from third most effective at α = 0.8)
- Remote counseling
- Healing ritual
- Emergency packages
- Teleworking

The order does not change at α = 0·4, but for α = 0·2 the order is:

- Door-to-door quick response group
- Mindfulness program, individual followup
- Face-to-face counseling
- Remote counseling (from fifth most effective at α = 0.6)
- Individual physical activity (from fourth most effective at α = 0.6)
- Healing ritual
- Emergency packages
- Teleworking

## 5 Conclusions and Recommendations

As seen in Table 2, various interventions can have different MEUs, even between ones with the same efficacy. For example, the difference between Teleworking (0·5876 MEU) and Healing ritual (0·6226 MEU) signals that the difference in nodes affected produce changes between their MEUs. This could be a result of religion being a big part of the lives of many Filipinos, as seen in the Philippine national motto: *Maka-Diyos, Maka-tao, Makakalikasan at Makabansa,* or translated into English: For God, For People, For Nature, And For Country (Rep. Act No. 8491, §40).^27^ Adding to that, the 2020 Census of Population and Housing conducted by the Philippine Statistics Authority shows that 78.8% of those surveyed were part of the Roman Catholic faith.^28^ Thus, this may be the reason why spirituality-based interventions could be effective.

One other variable that could affect the results is the number of nodes between affected nodes and the utility node. For example, a hypothetical intervention with a 60% efficacy to improve mental health would have more influence than another intervention with a 60% efficacy to improve spirituality (0·6510 MEU vs 0·3121 MEU), possibly because the effect of the second node is lessened by how many other nodes have to be influenced before the utility node is. This is also seen with another hypothetical intervention with the same efficacy to improve Access to Internet (0·3110 MEU), which has to affect more nodes than Spirituality before having an effect on the utility node.

This is the reason for using a weight on the interventions’ influence on the utility node. In Table 3, it is shown that the order of most effective interventions changes based on the weight α. For example, from Individual physical activity being the most effective intervention at α = 1, it is reduced to fifth most effective when α = 0.2. We also note that from this data, it can be concluded that interventions which directly address mental health are the most effective.

According to a paper by Tolk, et al., the complex and wide-reaching impact of the COVID-19 pandemic influenced various efforts to understand the implications on various countries’ populations. This was achieved by creating models that predicted the effect of the virus on different aspects of society, such as economic factors and the health of the population. However, such efforts required a collaborative effort between multiple disciplines, and were hindered by a lack of understanding between researchers.^29^ Therefore, this research could be a starting point for multidisciplinary studies to be conducted in the Philippines regarding the COVID-19 pandemic, and aid the development of frameworks for conducting multidisciplinary research.

Also to note is the implementation of these interventions. For many places in the Philippines, not all interventions could be considered due to pandemic restrictions and limited resources. It is still advised that the recommended interventions be observed in various areas in the Philippines as it would lend validity to the model and provide data toward improving it.

The study aims to become a framework for many more applications where networks are applied in a real-world setting. In a Bayesian network, implementing interventions would become simpler, and the effects of an intervention could even be studied on a specific subset of the population. Given enough accurate data, networks can aid greatly in terms of improving the quality of life for everyone.

## Data Availability

All data produced in the present study are available upon reasonable request to the authors

## Acknowledgements

This research is funded by Canada’s International Development Research Centre (IDRC) (Grant No. 109981-001) and by Canada’s International Development Research Centre (IDRC) and the UK Foreign, Commonwealth and Development Office (FCDO) (Grant No. 110554-001). This work is conducted under the banner of the Global South Artificial Intelligence for Pandemic and Epidemic Preparedness and Response Network (AI4PEP), which supports responsible, locally grounded AI innovations for public health preparedness and response across the Global South.

## Appendix A: Ideas of Drs. M. Teresa Tuason and C. Dominik Güss

### Other Potential Psychological Interventions during Pandemic

Clearly, the Covid-19 pandemic has affected mental health not only of Covid-19 patients but also of the general population in many countries. Probably not as much as needed, but many countries have offered mental health interventions for the population. Several scientific studies focused on testing the effectiveness of these psychological interventions with the goal to improve the mental health of people affected by Covid-19. With more and more studies showing that psychological interventions during Covid-19 have been effective to decrease anxiety and depression in different populations (e.g., Yang et al., 2020; Ye et al., 2022; Zhou et al., 2020), it was possible to conduct **meta-analyses** and integrate the findings of many studies on the effectiveness of these psychological interventions. One meta-analysis, for instance, studied the effectiveness of randomized controlled studies examining remote or online interventions for mental health during Covid-19 (Fischer-Grote et al., 2024). Results of the combined studies with over 8700 participants showed medium effects for anxiety and social functioning, and large effects for depression. However, no significant treatment effects were found for well-being, disordered eating, and psychological distress. Another meta-analysis including 10 studies investigated the effects of internet-based interventions for reducing stress, depression, and anxiety among university students during the COVID-19 pandemic (Malinauskas & Malinauskiene, 2022). All 10 studies analyzed showed significant effects of internet-based interventions for reducing stress and depression in university students. Anxiety was not reduced significantly. It is interesting to note that even meta-analyses do not come to the same conclusions – perhaps due to different samples. Whereas one meta-analysis found large effects on anxiety, the other one did not. One meta-analysis found large effect on stress, the other did not. Yet, both meta-analyses found strong treatment effects on depression.

Once the effectiveness of several psychological intervention programs during Covid-19 has been shown, the question arises **which type of intervention** is the most effective one. Research has shown that it is not a single one intervention, but often a set of combined interventions that is most beneficial. He et al. (2020), for example, reported a set of interventions in China during Covid-19 consisting of three parts: 24-hour hotline consultations, online video intervention with 45 episodes, and on-site crisis intervention sessions. A systematic review including 1,147 participants compared five different psychological interventions (Yang et al., 2021). The good news is, all interventions were effective. The most effective therapy was supportive therapy (ST) adjusted to the COVID-19-related mental crisis. SD was compared to behavioral therapy, traditional Chinese medicine therapy, nursing-based psychological therapy, and COVID-19-related standard training at reducing the anxiety-related symptoms.

One critical aspect of a psychological intervention is how it is tailored towards a specific target group, and some studies have concentrated on different **societal target groups.** Liu et al. (2021), for example, focused on university students and especially on their social isolation and loneliness due to classes switching from face-to-face to online mode of instruction. Inchausti et al. (2020) distinguished three different groups with different needs: healthcare workers engaged in the frontline, individuals who have been diagnosed with Covid-19 or have lost a family member or loved one due to Covid-19, and individuals with existing mental health conditions who are especially at risk. The authors did not further differentiate other groups in society, for example the general population with no current symptoms. Yet, they are also affected by social distancing, isolation, care for family members etc.

Based on existing literature on the effectiveness of psychological interventions during Covid-19, we are drawing the following conclusions:

1. Many psychological interventions have been effective, but, in general, the mental health needs of various societal groups affected by Covid-19 in various societies have been poorly handled (De Sousa et al., 2020; Duan & Zhu, 2020; Xiang et al., 2020).
2. It is not only one kind of intervention, but a combination of interventions that seems to be most promising for reducing anxiety, stress, and depressive symptoms during Covid-19. It must be noted that not all studies show consistent findings.
3. There is no “one-size-fits-all” kind of intervention. It is advantageous to adjust the kind of intervention (e.g., counseling telephone helplines, one-on-one online counseling, VR counseling) and the duration of the intervention to the specific target group (e.g., patients in intensive care unit, people at home, students), and to the specific location (e.g., intensive care unit, ordinary isolation ward, at home).
4. The Covid-19 pandemic, its effect on the people, and the resulting political policies change over time. Additionally, the symptoms of people affected by Covid-19 change over time. Thus, changes occur on the macro-societal and the micro-individual level. The pandemic and individuals’ suffering change. Psychological needs also change and with them interventions need to change. For example, even after being treated for Covid-19 and being released from hospital, most patients still show elevated symptoms of stress (PTSD) and need follow-up care.
5. We have not addressed here the political and practical aspects of psychological interventions. For any intervention to be effective, the context and the resources need to be identified. These include the political and practical aspects of the psychological interventions, the ways by which they are implemented, the extent of cooperation between national agencies and community health services and mental-health-care institutions, and the organizers of these interventions. In terms of resources, concerns about having enough resources, the mobilization of these resources, having enough experts such as psychologists, clinical psychiatrists, and mental health social workers, and having appropriate training for these interventions need to be addressed.
6. We would like to note that most studies have been conducted in China and the West, and not all results can be generalized to other countries and cultures. This is a big problem. Cultural values, for example, affect how politicians and societies react to the pandemic (Güss & Tuason, 2021). Thus, it is reasonable to assume that cultures also accept different forms of psychological interventions and react to specific forms of psychological interventions differently,

Thus, there is a need to further elaborate on psychological interventions during a pandemic and to consider the specific cultural context and specific cultural healing methods.

For the Filipino context, of course, interventions that may prove efficacious are the ones specific and relevant to the Philippines; and framed by both indigenizing movements such as knowledge, mental health practices and beliefs (Mijares, 2019) as well as those established in this Bayesian Network findings. We use the framework of the indigenous healing concepts identified by Enriquez (1977) championing *pakikipagkapwa* (being one with others), *pagmamalasakit* (concern), and *gaan nang loob* (rapport and acceptance), among others, and also indigenous healing that advocates for the natural way of being and being embodied within our communities and relationships through the interconnectedness of mind, body, and soul, and the use of local and folk remedies that honors the power of community activism and reaching those in the fringes of Philippine society (Zhou et. al., in press). Most especially, we utilized the nodes identified in the Bayesian network, summarized in Figures 1 and 2.

*<u>Kalusugang PangGrupo</u>*<u>: Group Mental Health Promotion.</u> Compared to many western approaches to healing focusing on the individual, the community-aspect is stronger and more important in the Filipino context. Activities that directly focus on taking care of mental health and well-being can be conducted through the media thru TV stations, radio stations, Zoom rooms, social media outlets or in open public spaces such as parks and barangay corners. These health promotion groups can be led by anyone with talent, expertise, or interest in any of the expressive areas and would foster not just empathy, accountability, and camaraderie, by being together, but they could directly promote wellness through natural healing practices such as expressive arts in dance (e.g., folk dance, hip hop) in physical activity (e.g., Tai-chi, Zumba, gymnastics, yoga), in meditation (e.g., nature-bathing, imagery), in singing (e.g., karaoke, chanting), in poetry (e.g., reading, writing), in spiritual practices (e.g., attending services, praying the rosary, novenas).

Group Mental Health Promotion would influence in Fig 1:

- Ease of access to resources:

○ Access to leisure/exercise
○ Access to social relationships
- Intrinsic properties

○ Sense of agency
- Direct impact of pandemic

○ Physical health
○ Loneliness
- Mental Health Indicators

○ Social relationships
○ Engaged in daily activities/competent and capable
○ Optimistic
- Mental Health

*<u>Bayanihan</u>*<u>: Community-Led Outdoor Interventions.</u> Projects that take place outdoors among families, in neighborhoods, in safe and open spaces can directly help with pursuing interests and leisure, while helping the community, in financial and well-being concerns. These activities can be led by mental health professionals and/or anyone in the neighborhood such as barangay leaders or young adult leaders. Participants can be recruited by word of mouth or through texting, and the groups can volunteer tasks where they have common interests such as community gardening, clean ups (e.g., after a typhoon), community food pantries, setting up medical, dental, or mental health missions, online consults for counseling and more importantly watching out for each other. During COVID-19 with social isolation in the Philippines, there sprung community pantries---places in the neighborhood where people bring surplus food to share with others. The community pantry is a way to share with society’s most vulnerable and a way of communal healing (Macaraan, 2022).

Community-Led Outdoor Intervention would influence in Fig 1:

- Ease of access to resources:

○ Access to social relationships
○ Access to healthcare
- Intrinsic properties

○ Sense of agency
○ Empathy
○ Socio-economic status
- Direct impact of pandemic

○ Loneliness
○ Financial blanket
- Mental Health Indicators

○ Purposeful and meaningful life/Good person, Good life
○ Contribute to well-being of others
○ Social relationships
○ Engaged in daily activities/competent and capable
○ Optimistic
○ Respected
- Mental Health

*<u>Katutubong Lunas</u>*<u>: Indigenous Healing Practices.</u> Western psychological interventions have heavily focused on cognitive and behavioral interventions. Only in recent years have Eastern concepts such as mindfulness been incorporated into Western psychology. For the Filipino context, cultural rituals and expressions that encourage and facilitate values of hope, faith, gratitude, healing, and belonging are essential. These activities (herbal medicine, visiting an *albularyo*, prayer, processing and asking questions *pagtatanong tanong*, storytelling *pakikipagkuwentuhan*) can be carried out individually in homes/families or in common spaces but can be scheduled at certain regular times so that barangay members can join in whenever they can, and community belongingness and sense of agency can be fostered. An example of this is *Dungaw,* where statues of saints or Mother Mary are put in places for others to see, and they promote hope and faith, and also deep connection with each other through care *pagdadamayan*, and affection *pagkalinga* (del Castillo et al., 2021).

Indigenous Healing Practices would influence in Fig 1:

- Ease of access to resources:

○ Access to social relationships
○ Access to healthcare
- Intrinsic properties

○ Sense of agency
○ Empathy
○ Spirituality
- Direct impact of pandemic

○ Loneliness
- Mental Health Indicators

○ Purposeful and meaningful life/Good person, Good life
○ Social relationships
○ Optimistic
- Mental Health

The three proposed interventions, *Kalusugang PangGrupo, Bayanihan, and Katutubong Pamana*, are inexpensive, and they are meant to be available to everyone in the barangays who is willing and open – regardless of socio-economic status. They tap into emerging leaders of the community, the established leaders in the barangays, or anyone who is available and willing to contribute their resources for the greater good. They can be mental health professionals, or they can be alternative healers *albularyos*, but most importantly they change the narratives of leadership in the Philippines, where the ultimate power is defined by helping the people in our communities who need it the most. Most importantly, all three champion a holistic approach to healing (Rondilla et al., 2021).

