## Supplementary material for "From the COVID-19 Pandemic to the Mental Health of the Philippines: Modelling the Cascade of Disasters using an Influence Diagram": Tables

Table 1: Efficacies and impacts of decision nodes.

| Decision Node | Efficacy | Impacts |
| --- | --- | --- |
| Mindfulness program, individual followup | 65% | Access to leisure/exercise, Access to social relationships, Sense of agency, Physical health, Loneliness, Social relationships, Engaged in daily activities/competent and capable, Optimistic, Mental health |
| Door-to-door quick response group | 65% | Access to social relationships, Sense of agency, Empathy, Socio-economic status, Loneliness, Purposeful and meaningful life, Contribute to well-being of others, Social relationships, Engaged in daily activities, Optimistic, Respected, Mental health |
| Healing ritual | 50% | Access to social relationships, Access to healthcare, Sense of agency, Empathy, Spirituality, Loneliness, Purposeful and meaningful life, Social relationships, Optimistic, Mental health |
| Face-to-face mental health counseling | 65% | Access to healthcare, Physical health, Empathy, Contribute to well-being of others, Optimistic, Loneliness, Living situation, Purposeful and meaningful life, Job security, Financial blanket, Access to social relationships, Mental health |
| Remote mental health counseling | 55% | Access to healthcare, Physical health, Empathy, Contribute to well-being of others, Optimistic, Loneliness, Living situation, Purposeful and meaningful life, Job security, Financial blanket, Access to social relationships, Access to internet, Sense of agency, Mental health |
| Individual physical activity | 75% | Physical health, Got infected by COVID, Activities, Sense of agency, Access to social relationships, Mental health |
| Teleworking | 50% | Got infected by COVID, Sense of agency, Socio-economic status, Loneliness, Access to internet, Online communication, Mental health |
| Emergency packages | 50% | Financial blanket, Contribute to well-being of others, Access to social relationships, Loneliness, Mental health |

Table 2: MEUs based on decision scenarios.

| Decision Node | Maximum Expected Utility if decision is ‘no’ (baseline MEU) | Maximum Expected Utility if decision is ‘yes’ |
|---|---|---|
| Individual physical activity | 0·3110 | 0·7905 |
| Mindfulness program, individual followup | 0·3110 | 0·7785 |
| Door-to-door quick response group | 0·3110 | 0·7595 |
| Face-to-face counseling | 0·3110 | 0·7467 |
| Remote counseling | 0·3110 | 0·6712 |
| Healing ritual | 0·3110 | 0·6226 |
| Emergency packages | 0·3110 | 0·6163 |
| Teleworking | 0·3110 | 0·5876 |

Table 3: MEUs based on decision scenarios, with the influence on Mental Health weighted.

| Decision Node | Maximum Expected Utility if decision is ‘yes’ ($\alpha = 1$, no change) | Maximum Expected Utility if decision is ‘yes’ ($\alpha = 0{\cdot}8$) | Maximum Expected Utility if decision is ‘yes’ ($\alpha = 0{\cdot}6$) | Maximum Expected Utility if decision is ‘yes’ ($\alpha = 0{\cdot}4$) | Maximum Expected Utility if decision is ‘yes’ ($\alpha = 0{\cdot}2$) |
|---|---|---|---|---|---|
| Individual physical activity | 0·7905 | 0·6819 | 0·5862 | 0·5033 | 0·4333 |
| Mindfulness program, individual followup | 0·7785 | 0·6913 | 0·6143 | 0·5476 | 0·4911 |
| Door-to-door quick response group | 0·7595 | 0·7014 | 0·6498 | 0·6047 | 0·5661 |
| Face-to-face counseling | 0·7467 | 0·6691 | 0·6004 | 0·5405 | 0·4896 |
| Remote counseling | 0·6712 | 0·6076 | 0·5507 | 0·5004 | 0·4567 |
| Healing ritual | 0·6226 | 0·5637 | 0·5105 | 0·4628 | 0·4206 |
| Emergency packages | 0·6163 | 0·5561 | 0·5016 | 0·4525 | 0·4091 |
| Teleworking | 0·5876 | 0·5231 | 0·4646 | 0·4121 | 0·3657 |
